# Age-Based Disparities in Hospitalizations and Mortality for Coronavirus Disease 2019 (COVID-19)

**DOI:** 10.1101/2021.06.14.21258919

**Authors:** Lauren E. Wisk, Santi K.M. Bhagat, Niraj Sharma

**Affiliations:** Division of General Internal Medicine and Health Services Research, David Geffen School of Medicine at the University of California, Los Angeles (UCLA); Physician-Parent Caregivers, Washington DC; Division of General Medicine, Department of Medicine, Brigham and Women’s Hospital, Boston, MA; Division of General Pediatrics, Department of Pediatrics, Boston Children’s Hospital, Boston, MA; Harvard Medical School, Boston, MA

**Keywords:** COVID-19, SARS-CoV-2, coronavirus, health inequities, chronic conditions, young adults, health equity, health disparities

## Abstract

**Purpose:** Evidence suggests that older adults, racial/ethnic minorities, and those with comorbidities all face elevated risk for morbidity and mortality from COVID-19; but there are limited reports describing the potential for interactions between these factors.

**Methods:** We sought to evaluate age-based heterogeneity in observed disparities in hospitalization, ICU admission, and mortality related to COVID-19 using CDC public use surveillance data on 3,662,325 COVID-19 cases reported from January 1 to August 30, 2020.

**Results:** Racial/ethnic and comorbidity disparities in hospitalization were most pronounced during ages 20-29 and ages 10-19, with similar elevation seen for disparities in ICU risk.

Racial/ethnic disparities in mortality were most pronounced during ages 20-29 while risk from comorbidity peaks among ages 10-39.

**Conclusions:** As COVID-19 continues to affect younger populations, special attention to the implications for the most vulnerable subgroups are clearly warranted.

**Implications and Contribution:** Adolescents and young adults appear to have experienced the greatest inequities in COVID-19 outcomes by race/ethnicity and comorbidity. Careful monitoring of trends in this population is warranted as they re-enter school, work, and social settings while being the last group to receive priority for vaccination.

## INTRODUCTION

The incidence of coronavirus disease 2019 (COVID-19) was highest for older adults early in the pandemic, but recent data highlights a changing age distribution such that during summer months, COVID-19 incidence was highest in persons aged 20–29 years, disproportionately accounting for >20% of all confirmed cases.^1^ Rising incidence in younger populations may be partially explained by the social, economic, occupational, and behavioral factors that both make this group unique and are known to increase risk for exposure to SARS-CoV-2.^1,2^ Moreover, younger populations are more racially/ethnically diverse than older populations and a significant number of young people currently live with chronic illnesses, like diabetes and asthma.^3^ Substantial disparities in COVID-19 outcomes have been documented by race/ethnicity and comorbidities.^4,5^ Yet, the impact of age on the magnitude of these disparities is under-characterized despite the potential to reveal important differences that may be masked by the presentation of age-adjusted vs age-specific analyses.^6^ Further, a health equity lens requires attention to specific peaks in risk among racial/ethnic minorities and those with comorbidities in order to best inform policies aimed at mitigating these disparities. As such, we sought to evaluate age-based moderation of observed disparities by race/ethnicity and comorbidities in hospitalization, ICU admission, and mortality related to COVID-19.

## METHODS

The Centers for Disease Control and Prevention (CDC) COVID-19 case surveillance system provides de-identified data on individual cases, including on demographics, comorbidities, and disease/clinical outcomes, as reported by US states and territories and other autonomous reporting entities via the CDC’s standard case reporting form.^4^ Per prior reports, individual-level case report data are available for approximately 68% of the aggregate number of confirmed cases.^1^ We obtained data on 3,662,325 patients from the dataset released in September 2020, which included cases reported between January 1 and August 30. American Community Survey data was used to determine population totals to estimate case rates per population.

In this cross-sectional analysis of case surveillance data, we calculated unadjusted relative risk of hospitalization, ICU admission, and mortality for racial/ethnic groups and comorbidity status by age group. Cases without a definitive outcome documented (e.g., 49% unknown hospitalization status) were treated as not having the outcome (yes vs no/unknown), consistent with prior reporting; a sensitivity analysis restricted analyses to cases with definitive outcomes (yes vs no). Negative binomial regression with interactions between age groups and race/ethnicity and comorbidity were used to examine outcomes while adjusting for time, sex, case status (confirmed vs probable), presence of symptoms, age, comorbidity, race/ethnicity.

## RESULTS

Of 3,662,325 cases, 9.4% were documented as hospitalized, 1.3% had a documented ICU admission, and 3.4% died (Table 1). Racial/ethnic disparities were more pronounced among ages 10-19 and 20-29 and tended to decline with age (Figure 1). Differences by comorbidity were also more pronounced in younger age groups and tended to decline with age (e.g., relative risk of mortality for those with vs without a comorbidity was 13.94, 95%CI:4.90-39.65, among ages 10-19 and 13.66, 95%CI:9.03-20.66, among ages 20-29). Within all age groups, disparities by comorbidity, and to a lesser extent race/ethnicity, were larger for mortality risk than for hospitalization or ICU admission. Findings were consistent after multivariate adjustment, when restricting to lab-confirmed cases only, and when restricting to those with definitive outcome status (data not shown).

**Table 1.** **Reported number of COVID-19 cases and estimated rate, by available demographics and clinical factors**

|  | Total Cases |  |  | Hospitalization |  |  | ICU Admission |  |  | Death |  |  |
| --- | --- | --- | --- | --- | --- | --- | --- | --- | --- | --- | --- | --- |
|  | N | % | Rate | N | % | Rate | N | % | Rate | N | % | Rate |
| **Total** | 3,662,325 |  | 1,149.9 | 342,768 | 9.4% | 107.6 | 46,274 | 1.3% | 14.5 | 125,870 | 3.4% | 39.5 |
| **Age Group** |  |  |  |  |  |  |  |  |  |  |  |  |
| 0-9 years | 108,383 | 3.0% | 274.6 | 2,434 | 2.2% | 6.2 | 276 | 0.3% | 0.7 | 44 | 0.0% | 0.1 |
| 10-19 years | 271,652 | 7.4% | 657.7 | 3,754 | 1.4% | 9.1 | 420 | 0.2% | 1.0 | 72 | 0.0% | 0.2 |
| 20-29 years | 684,226 | 18.7% | 1,575.1 | 16,309 | 2.4% | 37.5 | 1,459 | 0.2% | 3.4 | 562 | 0.1% | 1.3 |
| 30-39 years | 622,332 | 17.0% | 1,498.4 | 26,873 | 4.3% | 64.7 | 2,904 | 0.5% | 7.0 | 1,618 | 0.3% | 3.9 |
| 40-49 years | 580,660 | 15.9% | 1,445.7 | 38,266 | 6.6% | 95.3 | 5,182 | 0.9% | 12.9 | 3,973 | 0.7% | 9.9 |
| 50-59 years | 564,999 | 15.4% | 1,356.3 | 58,668 | 10.4% | 140.8 | 9,016 | 1.6% | 21.6 | 10,319 | 1.8% | 24.8 |
| 60-69 years | 396,925 | 10.8% | 1,055.3 | 70,019 | 17.6% | 186.2 | 11,344 | 2.9% | 30.2 | 21,262 | 5.4% | 56.5 |
| 70-79 years | 222,836 | 6.1% | 1,012.5 | 63,576 | 28.5% | 288.9 | 9,301 | 4.2% | 42.3 | 30,816 | 13.8% | 140.0 |
| 80+ years | 206,172 | 5.6% | 1,823.0 | 62,826 | 30.5% | 555.5 | 6,367 | 3.1% | 56.3 | 57,184 | 27.7% | 505.6 |
| Unknown | 4,140 | 0.1% | -- | 43 | 1.0% | -- | 5 | 0.1% | -- | 20 | 0.5% | -- |
| **Sex** |  |  |  |  |  |  |  |  |  |  |  |  |
| Female | 1,850,866 | 50.5% | 1,137.4 | 157,920 | 8.5% | 97.0 | 18,160 | 1.0% | 11.2 | 57,668 | 3.1% | 35.4 |
| Male | 1,713,591 | 46.8% | 1,100.0 | 179,632 | 10.5% | 115.3 | 28,018 | 1.6% | 18.0 | 67,207 | 3.9% | 43.1 |
| Other/Unknown | 97,868 | 2.7% | -- | 5,216 | 5.3% | -- | 96 | 0.1% | -- | 992 | 1.0% | -- |
| **Race/Ethnicity** |  |  |  |  |  |  |  |  |  |  |  |  |
| AIAN nH | 27,041 | 0.7% | 1,298.3 | 2,766 | 10.2% | 132.8 | 220 | 0.8% | 10.6 | 845 | 3.1% | 40.6 |
| Asian nH | 68,917 | 1.9% | 386.2 | 11,747 | 17.0% | 65.8 | 2,439 | 3.5% | 13.7 | 4,915 | 7.1% | 27.5 |
| Black nH | 412,907 | 11.3% | 1,068.2 | 69,291 | 16.8% | 179.3 | 9,264 | 2.2% | 24.0 | 24,153 | 5.8% | 62.5 |
| Hispanic/Latino | 622,825 | 17.0% | 1,065.0 | 67,361 | 10.8% | 115.2 | 9,268 | 1.5% | 15.8 | 18,359 | 2.9% | 31.4 |
| Multiple/Other nH | 92,105 | 2.5% | 1,047.2 | 12,575 | 13.7% | 143.0 | 1,614 | 1.8% | 18.4 | 4,602 | 5.0% | 52.3 |
| NHPI nH | 7,859 | 0.2% | 1,512.8 | 950 | 12.1% | 182.9 | 281 | 3.6% | 54.1 | 167 | 2.1% | 32.1 |
| White nH | 826,556 | 22.6% | 430.2 | 106,423 | 12.9% | 55.4 | 14,174 | 1.7% | 7.4 | 55,119 | 6.7% | 28.7 |
| Unknown | 1,604,115 | 43.8% | -- | 71,655 | 4.5% | -- | 9,014 | 0.6% | -- | 17,710 | 1.1% | -- |
| **Comorbidity** |  |  |  |  |  |  |  |  |  |  |  |  |
| No | 300,399 | 8.2% | -- | 13,228 | 4.4% | -- | 2,633 | 0.9% | -- | 1,795 | 0.6% | -- |
| Yes | 489,620 | 13.4% | -- | 143,275 | 29.3% | -- | 28,304 | 5.8% | -- | 62,052 | 12.7% | -- |
| Unknown | 2,872,306 | 78.4% | -- | 186,265 | 6.5% | -- | 15,337 | 0.5% | -- | 62,023 | 2.2% | -- |
| **Lab-confirmed case** | 3,521,190 | 96.1% | -- | 340,868 | 9.7% | -- | 46,134 | 1.3% | -- | 118,861 | 3.4% | -- |

AIAN, American Indian/Alaska Native; nH, non-Hispanic; NHPI, Native Hawaiian/Pacific Islander.
Column percentages are shown for total cases, row percentages are shown elsewhere.
Rates per 100,000 population calculated using the US Census Bureau's American Community Survey, 2017 & 2018, population estimates.

**Figure 1.**
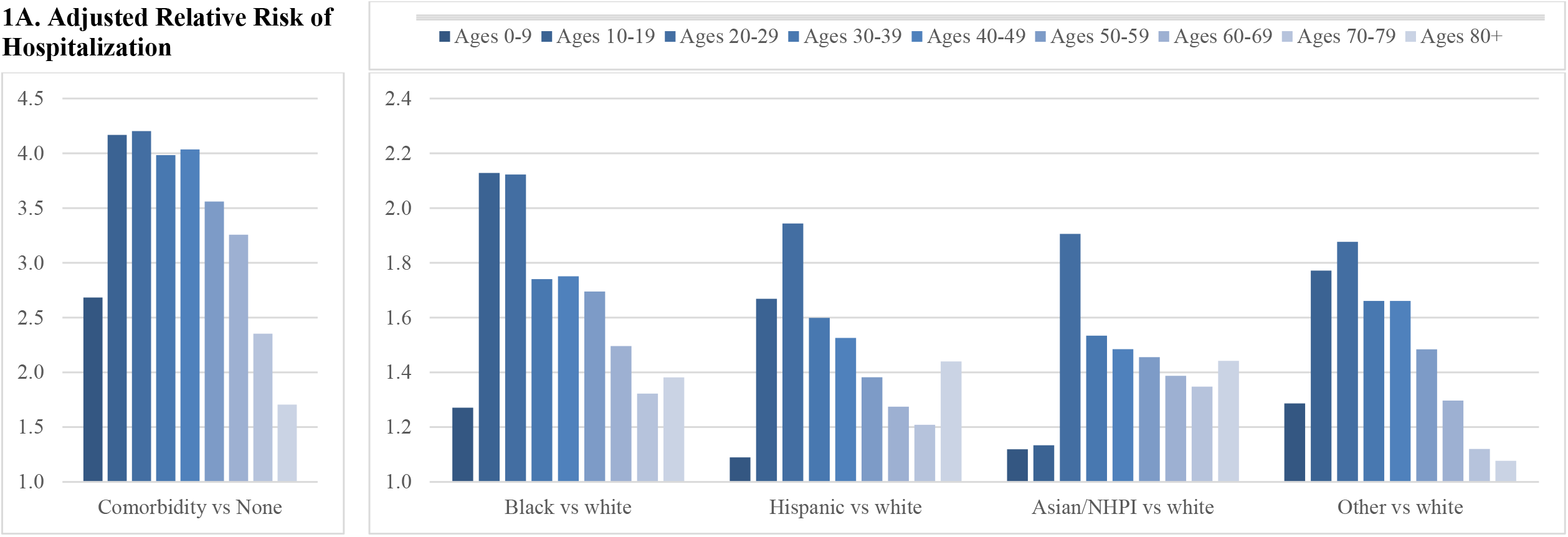
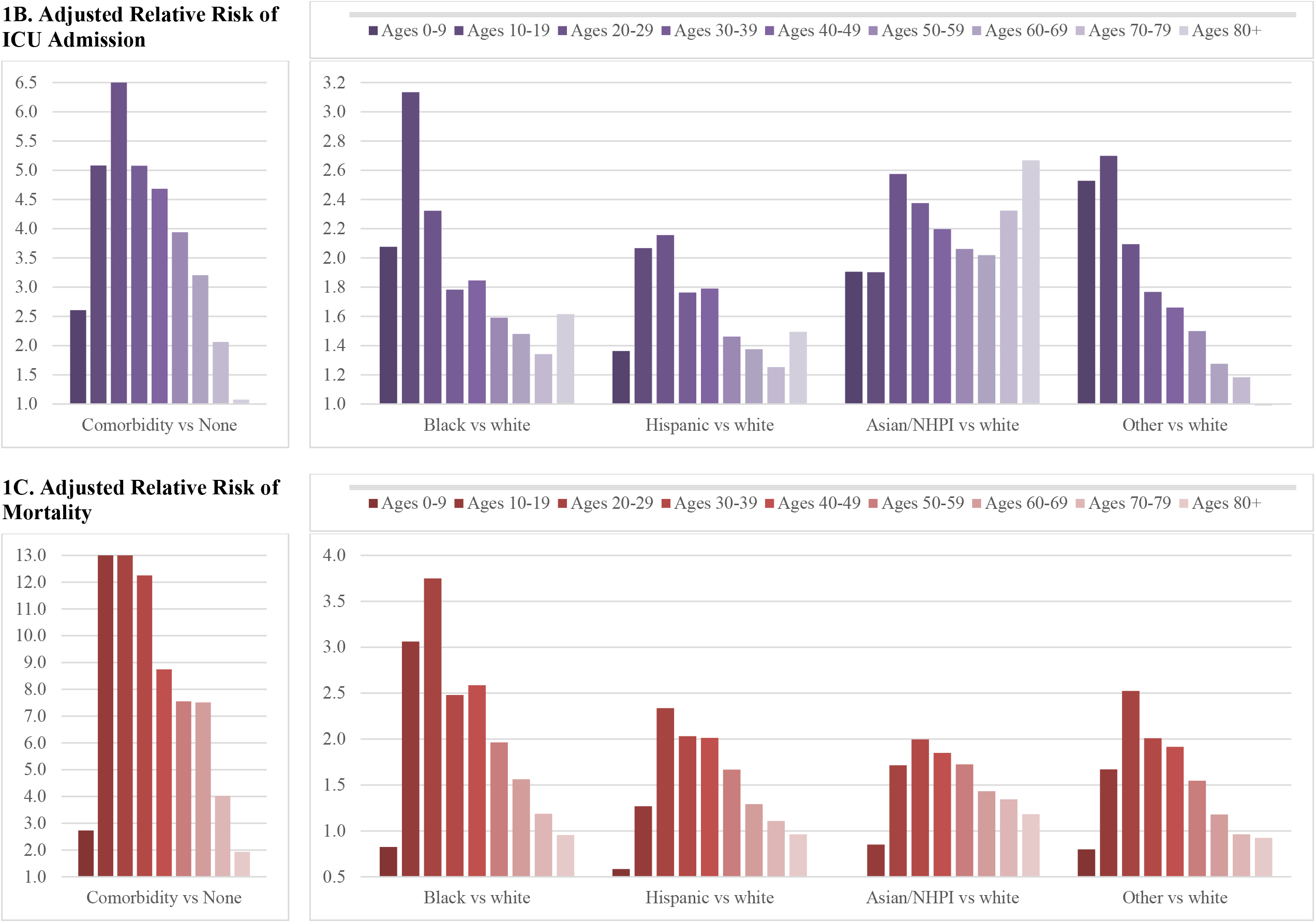
Age-based heterogeneity in the adjusted relative risk of COVID-19 outcomes by race/ethnicity and comorbidity. Figures show racial/ethnic and comorbidity-based disparities in the adjusted relative risk of hospitalizations (panel A), ICU admissions (panel B), and mortality (panel C) by age. Models adjust for time (week case was reported to the CDC), sex, case status (probable vs lab confirmed), whether case was symptomatic, age, comorbidity, and race/ethnicity, with interactions between age and comorbidity or race/ethnicity.

## DISCUSSION

Inequities in COVID-19 outcomes by race/ethnicity and comorbidity appear most pronounced for adolescents and young adults. Although absolute burden of COVID-19 is highest for older adults, a health equity lens requires attention to the relative elevation in risk among racial/ethnic minorities and those with comorbidities that peaks for a vulnerable younger population.^5^ Younger adults are a large proportion of essential workers,^1^ more likely to be un-/under-insured, less economically secure, more likely to experience unmet health care needs, and more likely to engage in risky behaviors^3^ – foreshadowing magnification of inequalities in COVID-19 outcomes during young adulthood. Yet, young adults are often under-recognized as distinct population, preventing a full understanding of their unique risks and needs.^3^ Given adverse outcomes among this population,^7^ and that poor health in young adulthood has important long-term consequences, this population warrants increased attention in public health, prevention, and equity efforts. Young adults likely need targeted messaging and guidance to address their potential role as disease vectors and their own susceptibility to COVID-19.^2^ Additional surveillance and research for this high-risk population are necessary as the pandemic continues and importantly, as younger populations are only just receiving access to vaccination.

Incomplete reporting by states/counties leads to considerable non-random missing data and lack of geographic coverage, likely limiting representativeness and potentially introducing measurement error.^8^ However, despite considerable limitations with CDC surveillance data, this dataset currently represents the best national accounting of COVID-19 cases in the US.

Inequalities are likely explained by residual confounding from root causes not captured by public-use surveillance data, such as socioeconomic disparities and the experience of systemic racism, and are thus proxied by the use of racial/ethnic groups (e.g., Black-white disparities are due to conscious and unconscious biases and discrimination that predispose Black individuals to worse outcomes and prohibit equitable treatment). The use of ten-year age brackets could mask patterns for subgroups, such as college students. Analyses only involve reported case data and cannot account for potential disparities in testing that could introduce selection biases. Finally, cases are temporally and geo-spatially evolving, thus findings may change over time and location.

## Data Availability

Data are publicly available from the CDC

## Abbreviations

COVID-19: Coronavirus disease 2019
CDC: Centers for Disease Control and Prevention
RR: Relative risk
CI: Confidence interval
ICU: Intensive care unit

## Acknowledgements

None

